# Sexuality in female patients on hemodialysis: a scoping review protocol

**DOI:** 10.1101/2024.08.15.24311979

**Authors:** Solange Imhof, Fabiana Baggio Nerbass, Rafaela Gonzaga dos Santos, Heitor S. Ribeiro, Luana Gabriele Nilson

**Affiliations:** Centro de Tratamento de Doenças Renais, Jaraguá do Sul, Santa Catarina, Brasil; Programa de Pós-Graduação em Saúde Coletiva - Universidade Regional de Blumenau – Blumenau, Brasil; Fundação Pró-Rim, Joinville – Santa Catarina, Brasil; Fundação Pró-Rim, Jaraguá do Sul –Santa Catarina, Brasil; LIM 12, Hospital das Clínicas HCFMUSP, Faculdade de Medicina, Universidade de São Paulo, São Paulo - São Paulo, Brasil

**Author notes:** **CORRESPONDING AUTHOR:** Solange Imhof.

**Keywords:** sexuality, hemodialysis, women

## Abstract

Sexuality is a human biological function that is not limited only to genitality but to total corporeality. Sexual function is a complex biopsychosocial phenomenon in which internal and external stimuli are modulated by the central and peripheral nervous system, resulting in biochemical, hormonal, and circulatory changes that culminate in physical and cognitive sexual results. Several conditions influence sexuality, and one of the diseases that can impair female sexual function and impact sexuality is chronic kidney disease. Considering the complexity and specificity of chronic kidney disease, the significant repercussions of hemodialysis treatment on the lives of patients, and the scarce scientific production, the need to delve deeper into the topic was perceived since the experience of sexuality of these women is relevant in the quality of life and not usually addressed by health professionals. To investigate the topic in-depth, a scoping review will be developed on the sexuality of female people undergoing hemodialysis treatment, regardless of their sexual orientation. The objective of this work is to map the production of knowledge about sexuality in women undergoing hemodialysis treatment. The search for publications will be carried out in the following databases and bibliographic index: Medical Literature Analysis and Retrieval System Online (MEDLINE via PubMed), Web of Science, in the Bibliographic Index of Latin American and Caribbean Literature in Health Sciences (LILACS) and APA PsycNet. In addition to these databases, the research will also include gray literature, such as Google, Google Scholar, and YouTube. The question that will guide the review is: “What is the current knowledge about sexuality in women on hemodialysis? The review will consider studies that include the terms “sexuality”, “women” and “hemodialysis”, without restrictions on language and date.

**Transfers:** Download data is not yet available.

**Metrics:** No metrics found.

## 1. INTRODUCTION

Sexuality is a human biological function that is not limited only to genitality but to total corporeality (Vieira, Souza, Nakamura & Mattar, 2012). Sexual function is a complex biopsychosocial phenomenon in which internal and external stimuli are modulated by the central and peripheral nervous system, resulting in biochemical, hormonal, and circulatory changes that culminate in physical and cognitive sexual results. Nowadays, human sexual behavior transcends the reproductive aspect and has come to be considered an integral part of the quality of life (Vieira, Souza, Nakamura & Mattar, 2012). Several conditions influence sexuality. Non-communicable chronic diseases, such as chronic kidney disease, diabetes mellitus, cardiovascular diseases, hypo/hyperthyroidism, cancer, and liver failure may harm female sexual function and lead to the development of a dysfunctional condition (Abdo, 2014). Chronic kidney disease is a condition that affects patients and their families in all aspects of life. One of them scarcely explored and which has a direct relationship with quality of life is sexuality in patients on dialysis (Ramirez, 2018). Sexual dysfunction disorders are not uncommon in people on dialysis. Reducedlibido affects women more than men (Munoz, 2010).

According to the World Health Organization, “in recent years, international human rights instruments have been increasingly used to support and advance legal claims by individuals and whole communities so that national governments will guarantee the respect, protection and fulfilment of their sexual and reproductive health rights (Cook et al., 2003)”.

Maintaining sexual health is a lifelong process with important implications far beyond the reproductive years. More work is needed to elaborate and develop interventions and guidance for improving sexual health and well-being across the life-course that go beyond a focus on specific problems, such as sexually transmitted infections or sexual violence. HRP will continue to research in this area, says Lale Say (WHO, 2023).

The literature does not show uniformity in the understanding of sexuality, and there is a lack of scoping reviews addressing the sexuality of women who experience kidney disease and require treatments such as hemodialysis. Furthermore, no official documents highlight the importance of researching this topic or even addressing women’s sexuality in a comprehensive, inclusive way that is not focused on reproduction or injury prevention.

The present study will be a scoping review of the sexuality of female people on chronic HD treatment, regardless of their sexual orientation. Considering the complexity and specificity of chronic kidney disease, the significant repercussions of hemodialysis treatment on the lives of patients, and the scarce scientific production, there is a need to delve deeper into the topic since the experience of sexuality of these women is relevant in the quality of life and not usually addressed by healthcare professionals.

### 1.1 Object of study

The object of study is to map the knowledge about the sexuality of women on hemodialysis.

## 2. METHODS

### 2.1 Type, period, and study setting

This is a scoping review, which is a form of knowledge synthesis that addresses a research question and allows the mapping of key concepts, types of evidence, and gaps related to a topic, carrying out analyses of primary studies with the potential to develop new research strategies for the problem discussed (Peters, 2021). The research protocol will be registered by the main author at Scielo as a pre-print. The five steps recommended by the Joanna Briggs Institute will be adopted: identification of the research question, search for relevant studies, study selection, data mapping, grouping, summarizing, and presenting results. Three Boolean operators will be used: “AND” and “OR” and “NOT” to restrict unnecessary evidence, in the following databases and bibliographic index: Medical Literature Analysis and Retrieval System Online (MEDLINE via PubMed), Web of Science, in the Bibliographic Index of Latin American and Caribbean Literature in Health Sciences (LILACS) and APA PsycNet. The choice of these databases and bibliographic index is due to the large number of studies in the healthcare area. In addition to these databases, the research will include gray literature, such as Google, Google Scholar, and YouTube, in January 2024. The proposed scoping review will beconducted under the JBI Methodology for Scoping Reviews and the Preferred Reporting Items for Systematic Reviews and Meta-Analyses for Scoping Reviews (PRISMA-ScR) extension.

The JBI Scoping Review Network is an international evidence-based healthcare organization of methodologists that develops resources and instructs individuals, organizations, and institutions on scoping review approaches. The JBI Handbook for Evidence Synthesis guides authors who wish to conduct systematic and scoping reviews following JBI methodologies (JBI Scoping Review Network).

Prisma also provides information on reporting guidelines designed to help authors report transparently why their systematic review was conducted, what methods they used, and what they found (PRISMA Extension for Scoping Reviews). The main PRISMA reporting guideline guides reporting systematic reviews that evaluate the effects of interventions and for different types or aspects of systematic reviews and other types of evidence synthesis, e.g., scoping reviews (PRISMA).

### 2.2 Eligibility criteria

The question that will guide the review is: “What is the production of knowledge in the literature about sexuality in women on hemodialysis?” To answer it, the review will consider all studies that explore “sexuality, women and hemodialysis”, without restrictions on language and date (**Table 1**). This scoping review will consider evidence from articles/studies that describe the sexuality of female adults with CKD (≥18 years) on hemodialysis. Evidence that includes other kidney replacement therapies (e.g., peritoneal dialysis and kidney transplantation), predialysis stage, pediatric patients (<18 years old), or those affected by acute kidney injury will not be included. “Woman” will be considered “female or female human being, endowed with intelligence and articulated language, bipedal, two-legged, classified as a mammal of the primate family, with the characteristic of an upright position and considerable size and weight of the body skull” (online Portuguese dictionary).

**Table 1.**
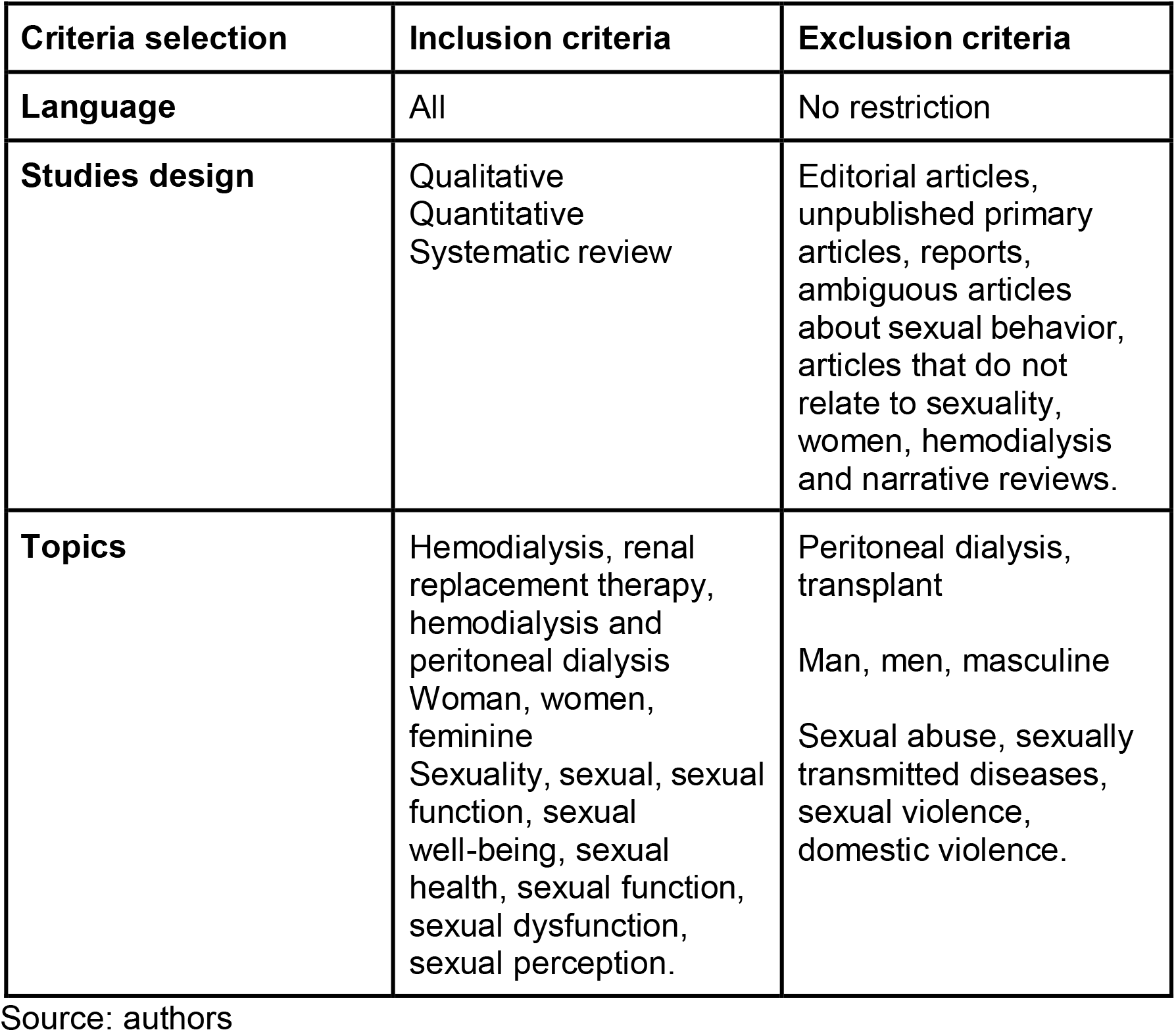
Criteria selection.

### 2.3 Context

No context restrictions will be applied to the project. Evidence from all available environments that address the sexuality of women undergoing hemodialysis treatment will be considered.

### 2.4 Types of fonts

This scoping review will consider all types of study and evidence (e.g., experimental, quasi-experimental, observational, etc.). Additionally, gray literature sources, websites, books, and guidelines from important societies and associations will be researched. On the other hand, systematic reviews, conference abstracts, comments, and letters to the editor will not be included. Evidence such as websites, books, and guidelines from important societies and associations that have described the sexuality of women on hemodialysis will also be considered.

### 2.5 Assessment of methodological quality

The same reviewers (S.I and F.B.N) will analyze the selected articles independently, considering the inclusion and exclusion criteria mentioned above. Questions are focused on key concepts related to the study, research question, and eligibility. For example, the clarity of the objective, the definition, and the appropriate description of exposure and outcomes will be considered essential for the study’s methodological quality.

### 2.6 Search strategy

The search strategy will aim to locate published peer-reviewed studies and non-scientific evidence. The text words contained in the titles and abstracts of the relevant articles and the index terms used to describe the articles will be used to develop a comprehensive search strategy in the databases and bibliographic index: Medical Literature Analysis and Retrieval System Online (MEDLINE via PubMed), Web of Science, in the Bibliographic Index of Latin American and Caribbean Literature in Health Sciences (LILACS) and APA PsycNet. The search strategy, including all identified keywords and index terms, will be adapted for each database and/or information source included. The reference list of all included evidence sources will be examined for additional studies. No language or date restrictions will apply. Gray literature sources will include Google Scholar, Google.com, and YouTube. The keywords used for the exhibition of interest will be Sexuality AND Women AND “Renal Dialysis” OR “Continuous Renal Replacement Therapy”, in combinations to suit the different databases, with the search keys described in Table 2.

**Table 2.**
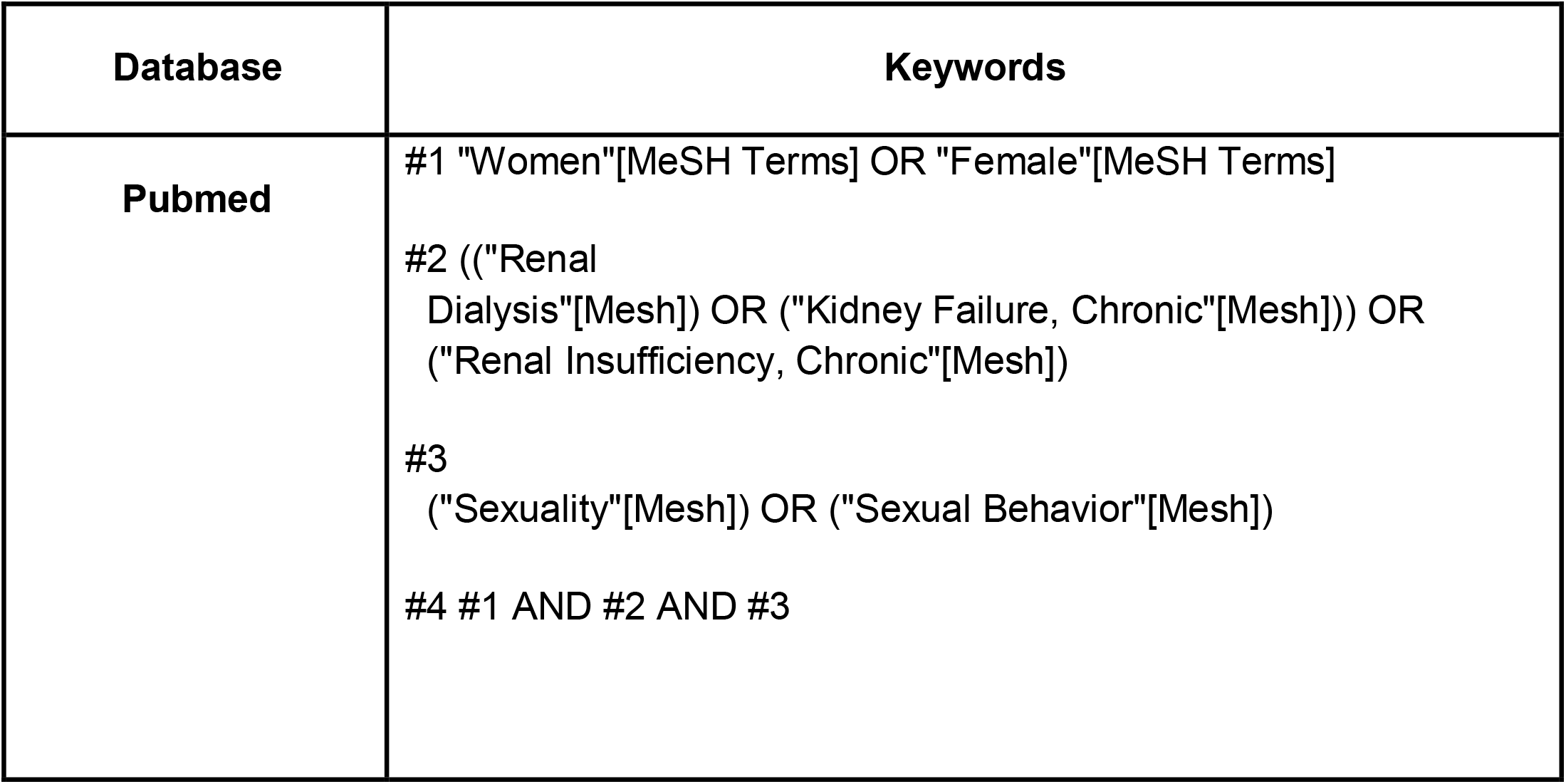
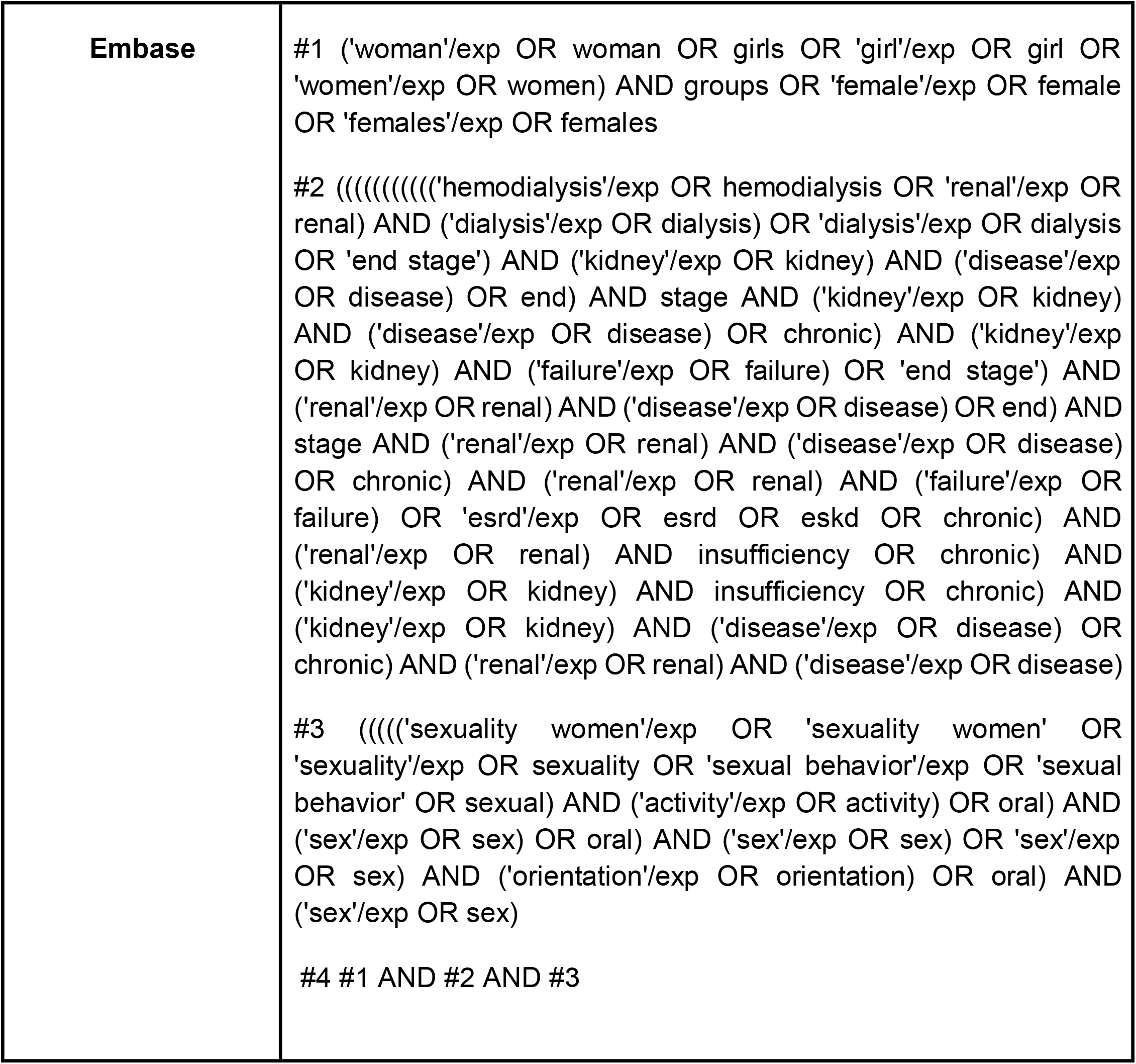
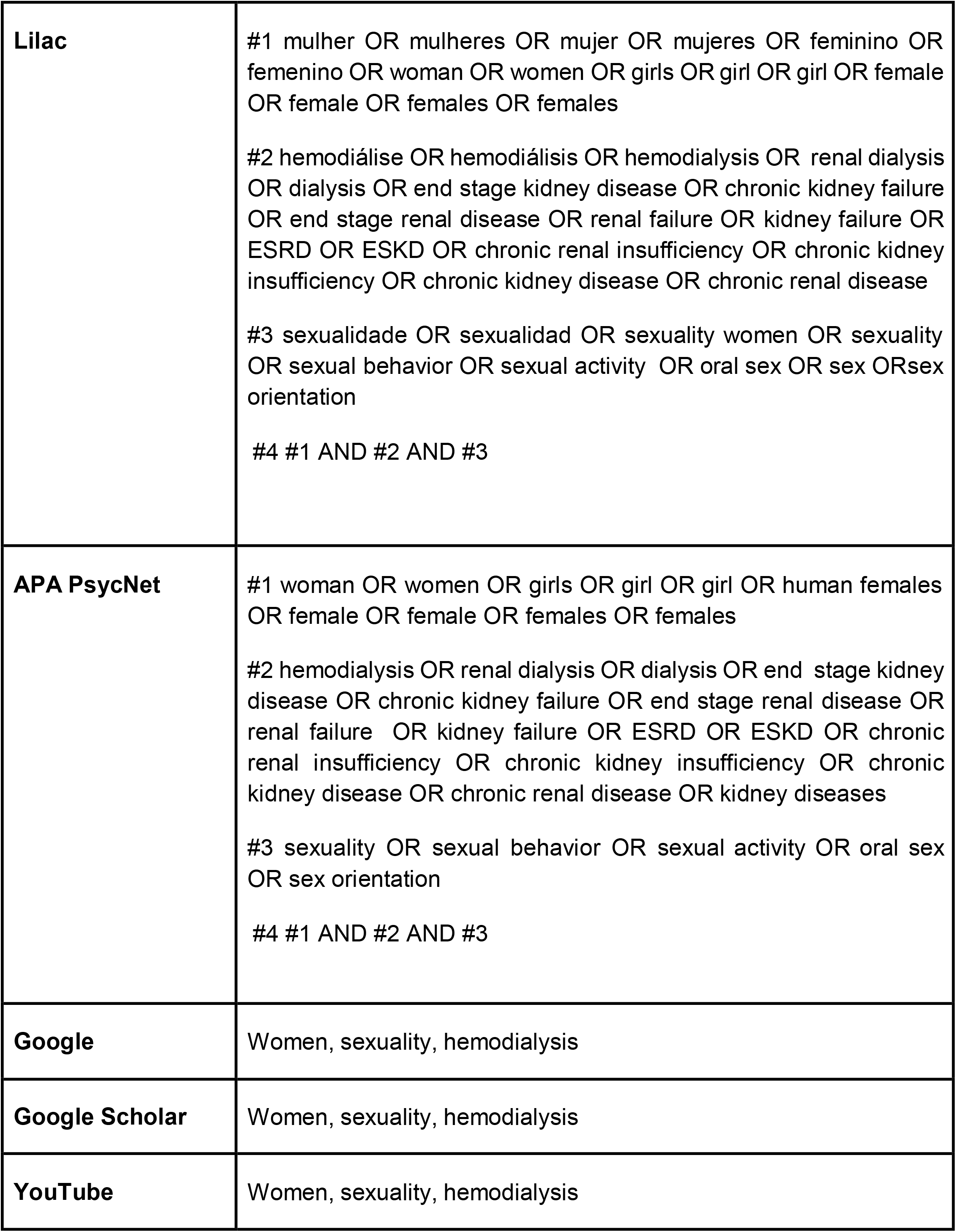
Search strategy according to database and total of references.

### 2.7 Selection of the source of evidence

After the search, references will be collated and loaded into the Covidence software (Veritas Health Innovation, Melbourne, AU), and duplicates will be removed for the screening process through the EndNote Web software. After a pilot test, two independent reviewers (S.I. and F.B.N) will examine the titles and abstracts for assessment according to inclusion criteria. The full text of the selected evidence will be assessed in detail according to the inclusion criteria by the same two independent reviewers. Reasons for excluding full-text sources of evidence that do not meet the inclusion criteria will be recorded and reported in the scoping review. The search and study inclusion process results will be reported in full in the final scoping review and presented in the PRISMA-ScR flow diagram.

### 2.8 Data analysis and presentation

The qualitative and quantitative results will be summarized and presented in tables and figures, with a narrative summary. The frequencies of the extracted data will be displayed in network diagrams and/or heat maps. A narrative summary will accompany the illustrations and describe how the results relate to the review objective and questions. The results of the scoping review will be reported in accordance with the PRISMA-ScR14 guideline. The categories will be analyzed based on the following themes: sexuality summarized as sexual dysfunction, depression, and changes in quality of life impact sexuality, sexuality, and the perception of oneself as a social construction.

### 2.9 Ethical aspects

As it is a scoping review, the project does not need to be submitted to the Research Ethics Committee, but it will follow all the rigor for preparing the research protocol.

## Data Availability

All data produced in the present study are available upon reasonable request to the authors.

## Authors’ contribution statement

SI: Conceptualization, data curation,investigation, visualization,writing – original draft, writing – review & editing.

FBN: Conceptualization, data curation,investigation, writing – original draft, writing – review & editing.

RGS:investigation,writing – review & editing.

HSR: Conceptualization, methodology, project administration,supervision, writing – review & editing.

LGN: Conceptualization, methodology, project administration,supervision, writing – review & editing.

## Research data availability statement

The entire dataset supporting the results of this study was published in the article itself.

